# Pharmaceutical Payments to Japanese Certificated Hematologists: A Retrospective Analysis of Personal Payments from Pharmaceutical Companies between 2016 and 2019

**DOI:** 10.1101/2022.01.17.22269284

**Authors:** Eiji Kusumi, Anju Murayama, Sae Kamamoto, Moe Kawashima, Makoto Yoshida, Hiroaki Saito, Toyoaki Sawano, Erika Yamashita, Tetsuya Tanimoto, Akihiko Ozaki

## Abstract

**Background:** A growing and significant financial relationship exist between physicians and pharmaceutical companies. However, little is known about the characteristics and trends of personal payments from pharmaceutical companies to hematologists. This study was aimed to evaluate the financial relationship between hematology specialists and pharmaceutical companies in Japan between 2016 and 2019.

**Methods:** Descriptive analyses were performed to evaluate personal payments from 92 major pharmaceutical companies to all board-certificated hematologists in Japan. Furthermore, trend of payments over four years were evaluated by generalized estimating equations.

**Results:** Among the 4,183 hematology specialists, 2,706 (64.7%) received a total of US$36,291,434 (¥3,955,766,292). The personal payments were worth $13,411 (standard deviation: $34,856) on average, with a median of $2,471 (Interquartile range: $851 $9,677) over the four-year period, respectively. Only the top 10% of specialists accounted for 76.8% of the total payments. The average payment values constantly increased from $4,259 to $5,574 between 2016 and 2019, with a significant mean annual increase rate of 11.2% (95%CI: 9.1% 13.4%). The proportion of hematology specialists with payments also slightly increased by 1.8% (95%CI: 0.6% 3.0%) per year.

**Conclusions:** Most Japanese board-certified hematology specialists received substantial personal payments. These payments are becoming increasingly more prevalent and greater among hematology specialists.

## Introduction

Although collaboration between healthcare professionals, patients, and pharmaceutical companies has played a significant role in the development of novel drugs and a better understanding of diseases (ref.[1, 2]), proper management of conflicts of interest (COIs) is currently one of the most fundamental concerns of all healthcare professionals (ref.[3]), as financial COIs between healthcare sectors and pharmaceutical companies could jeopardize patient-centered care. Indeed, the financial relationship between the healthcare sector and pharmaceutical companies might affect physician prescribing patterns (ref.[4–6]), recommendations in clinical guidelines (ref.[7–11]), and conducting and reporting of clinical research (ref.[12, 13]). These concerns and motivation to increase transparency in financial COIs has led to the creation of transparency initiatives worldwide (ref.[14, 15]), including in Japan. The Japan Pharmaceutical Manufacturers Association (JPMA), the largest pharmaceutical trade organization in Japan, requires all JPMA member companies from 2013 onwards to voluntarily publish all payments made to healthcare professionals and healthcare organizations (ref.[16]).

In the field of hematology, a wide variety of therapeutic strategies and novel therapies (ref.[17–19]), including chemotherapy, targeted therapy, and hematopoietic stem cell transplantation for the treatment of hematological cancers, have brought significant therapeutic benefits to patients and attracted considerable attention from pharmaceutical companies, resulting in intense marketing by pharmaceutical companies. The financial relationships of hematologists and oncologists with pharmaceutical companies as compared to other specialists have increased in recent years (ref.[20–22]). In addition, Japanese hematology guideline authors receive an average of $25,471 to $49,693 in personal payments from pharmaceutical companies (ref.[23]), one of the largest average payments among guideline authors in several specialties (ref.[11, 24, 25]). Considering the current pharmaceutical marketing strategy and the sizable evidence indicating that pharmaceutical marketing could influence physician prescription patterns (ref.[5, 26, 27]), the elucidation and proper management of the financial relationships between pharmaceutical companies and hematologists are important.

This study aims to elucidate the prevalence of board-certified hematologists receiving payments from pharmaceutical companies, the magnitude of the payments, and the payment trend in recent years.

## Methods

### Participants

This study considered all hematologists certified by the Japanese Society of Hematology, as of October 10, 2021, as hematology specialists. The Japanese Society of Hematology was established in 1937 and is the most prestigious and largest medical professional society in hematology in Japan, with 7,744 members as of October 8, 2021. The Japanese Society of Hematology is the sole professional medical society authorized to certify hematology specialists in Japan. The list of names of the hematology specialists is publicly available on the society’s webpage and was last updated on July 7, 2021 (http://www.jshem.or.jp/modules/senmoni/).

### Certification of hematologists by the Japanese Society of Hematology

The Japanese Society of Hematology is the sole society in Japan authorized to certify hematology specialists. As of October 2021, to be certificated as a hematology specialist, physicians are required to meet all of the following six requirements: (1) submission of clinical records for 15 inpatients as an attending physician, including at least three cases of erythrocyte disease, three cases of leukocyte disease, two cases of thrombosis and hemostasis, and one case of immunology and blood transfusion; (2) must be listed as the first author of at least two academic articles or conference presentations; (3) must have completed at least six years of clinical practice training after having acquired a medical license and at least three years of specialized training in hematology at an institution accredited by the Japanese Society of Hematology; (4) must have a board certification in internal medicine issued by the Japanese Society of Internal Medicine or in pediatrics issued by the Japan Pediatric Society; (5) must be a registered member of the Japanese Society of Hematology for at least three years; and (6) must clear the written examination for the hematology specialists conducted by the Japanese Society of Hematology.

### Data collection

The names, affiliations, and addresses of all hematology specialists were disclosed and collected from the Japanese Society of Hematology webpage on October 10, 2021. Payment data of all healthcare professionals and healthcare organizations regarding lecturing, writing, and consulting were collected for the period 2016-2019 from 92 pharmaceutical companies belonging to the JPMA. After collecting the payment data, a single unified database was developed, as described previously (ref.[24, 28]). JPMA requires the member companies to disclose only the payment concerning lecturing, writing, and consulting, along with the individuals’ names and affiliations; therefore, these payment categories could be analyzed on an individual specialist basis. Personal payments, such as those for lecturing, writing, and consulting, are directly and widely paid to specialists by pharmaceutical companies. (ref.[6, 28–31]) Considering the nature of payments, only the personal payments concerning lecturing, writing, and consulting were included in this study.

We scanned for specialist names in the payment database and extracted the payment data from the payment database of hematology specialists. The extracted data included the recipient names, affiliations, monetary amount, number of payment cases, payment category, and names of the respective pharmaceutical companies. To remove payment data of different persons with similar names in the database, we checked and compared the affiliations, affiliation addresses, and recipient specialties between the data from the Japanese Society of Hematology and the pharmaceutical companies For payments to specialists whose affiliations reported by the pharmaceutical company differed from those reported by the society, we manually searched the name of the specialists on the internet and collated other data from official institutional webpages and other sources to verify their identity. We also excluded payments that could not be verified from the analysis. The detailed procedure has been previously described (ref.[24, 28]). Furthermore, the lists of drugs newly approved for hematological diseases between 2015 and 2020 were collected from the webpage of the Pharmaceuticals and Medical Devices Agency, which is the official agency for reviewing drugs in Japan.

### Analysis

Descriptive analyses of payment values and the number of cases were performed per specialist and per pharmaceutical company. Average and median payments, cases, and number of companies making payments per specialist were calculated based only on the number of specialists receiving payments each year, as in other studies. (ref.[20, 29, 30, 32]) As several companies such as Shire Japan and Baxalta did not disclose the number of payment cases, the payments from these companies were excluded from the analysis of the number of cases, while the payment values were included in the analysis. To examine the concentration of payments to individual specialists, the Gini index and shares of the value of payments held by the top 1%, 5%, 10%, and 25% of specialists were calculated. The Gini index ranges from 0 to 1, and the greater the Gini index, the greater the disparity in the distribution of payments on a specialist basis, as mentioned previously. (ref.[24, 25]) Payment distribution was also examined geographically.

Further, to examine the trend of payments from pharmaceutical companies to specialists from 2016 to 2019, the population-averaged generalized estimating equation negative binomial regression model for the trend of payment value and the linear generalized estimating equation model log linked with binomial distribution for the trend of numbers of specialists with payments were used. The relative ratios of the average annual increase in payments per specialist to the number of specialists with payments were used to report the results.

The year of payments was set as an independent variable, and the proportion of physicians receiving payments, number of payments, and payment values were set as dependent variables. (ref.[20, 33, 34]) As several pharmaceutical companies disaffiliated from the JPMA and newly joined the JPMA, among all 92 companies, there were 18 companies without payment data over the four years. Thus, the average and median payments for each year and the trend of payments were calculated based on payments from all 92 companies and 74 companies with payment data for the four years between 2016 and 2019.

To adjust for inflation and to make the payment value comparable, Japanese yen (¥) was converted into US dollars ($) using the 2019 average monthly exchange rate of ¥109.0 per $1. All analyses were conducted using Microsoft Excel (version 16.0; Microsoft Corp.) and Stata (version 15; StataCorp).

### Ethical approval

This study was approved by the Ethics Committee of the Medical Governance Research Institute. As this study is a retrospective analysis of publicly available information, informed consent was waived and direct contact with the society was allowed by the Ethics Committee of the Medical Governance Research Institute.

## Results

### Overview of payments

We identified 4,183 hematology specialists certified by the Japanese Society of Hematology as of October 10, 2021. Our payment database recorded 1,474,653 payment cases and $996,291,009 (¥108,595,720,027) in total monetary value from 92 pharmaceutical companies between 2016 and 2019.

Of the 4,183 specialists, 2,706 (64.7%) received a total of US$36,291,434 (¥3,955,766,292) corresponding to 47,863 cases from 71 (77.2%) pharmaceutical companies between 2016 and 2019 (Table 1). The payment cases and values to the hematology specialists occupied 3.2% and 3.6% of total cases and values, respectively. A total of 82.5% ($29,951,526) of the total payments were for lecturing, followed by 13.5% from consulting ($4,890,255), 3.9% from writing ($1,398,729), and 0.1% from others ($50,924). The average monetary value per case was $758, ranging from $736 for lecturing to $922 for consulting.

**Table 1.** Summary of personal payments from pharmaceutical companies to hematology specialists certified by the Japanese Society of Hematology between 2016 and 2019

| **Variables** |  |
| --- | --- |
| **Total** |  |
| **Payment values, US\$ (%)** | **36,291,434 (3.6)** |
| **Cases, n (%)** | **47,863 (3.2)** |
| **Companies, n (%)** | **71 (77.2)** |
| **Average per specialist (SD)** |  |
| **Payment values, US\$** | **13,411 (34,856)** |
| **Cases, n** | **17.7 (35.9)** |
| **Companies, n** | **5.5 (5.2)** |
| **Median per specialist (IQR)** |  |
| **Payment values, US\$** | **2,471 (851□9,677)** |
| **Cases, n** | **5 (2□17)** |
| **Companies, n** | **4 (2□8)** |
| **Range** |  |
| **Payment values, US\$** | **46□528,038** |
| **Cases, n** | **0□487** |
| **Companies, n** | **0□32** |
| **Physicians with specific payments, n (%)** |  |
| **Any payments** | **2,706 (64.7)** |
| **Payments >US\$500** | **2,392 (57.2)** |
| **Payments >US\$1,000** | **1,947 (46.6)** |
| **Payments >US\$5,000** | **980 (23.4)** |
| **Payments >US\$10,000** | **666 (15.9)** |
| **Payments >US\$50,000** | **175 (4.2)** |
| **Payments >US\$100,000** | **78 (1.9)** |
| **Gini index** | **0.856** |
| **Category of payments** |  |
| **Lecturing** |  |
| **Payment value, US\$ (%)** | **29,951,526 (82.5)** |
| **Cases, n (%)** | **40,686 (85.0)** |
| **Consulting** |  |
| **Payment value, US\$ (%)** | **4,890,255 (13.5)** |
| **Cases, n (%)** | **5,302 (11.1)** |
| **Writing** |  |
| **Payment value, US\$ (%)** | **1,398,729 (3.9)** |
| **Cases, n (%)** | **1,816 (3.8)** |
| **Other** |  |
| **Payment value, US\$ (%)** | **50,924 (0.1)** |
| **Cases, n (%)** | **59 (0.1)** |

### Payments per specialists

The hematology specialists received personal payments worth $13,411 (Standard deviation (SD): $34,856) on average and a median of $2,471 (Interquartile range (IQR): $851 $9,677) over the four-year period. The average and median number of cases over the four years was 17.7 (SD: 35.9) and 5.0 (IQR:2.0 17.0) cases per specialist, respectively. The average number of companies making payments per specialist was 5.5 (SD: 5.2; median: 4.0; IQR: 2.0 8.0; and range: 1.0 32.0) (Table 1).

For the concentration of payments, 4.2% and 1.9% of hematology specialists received more than $50,000 and $100,000, respectively. The Gini index for the four-year cumulative payments per specialist was 0.856, indicating that the distribution of the total payments per specialist was highly skewed. Top 1%, 5%, 10% and 25% of specialists occupied 26.0% (95% confidence interval (CI): 23.1% 28.9%), 61.0% (95%CI: 58.2% 63.7%), 76.8% (95%CI: 74.8% 78.8%), and 93.3% (95%CI: 92.6% 94.0%) of total payments, respectively (Figure 1).

**Figure 1.**
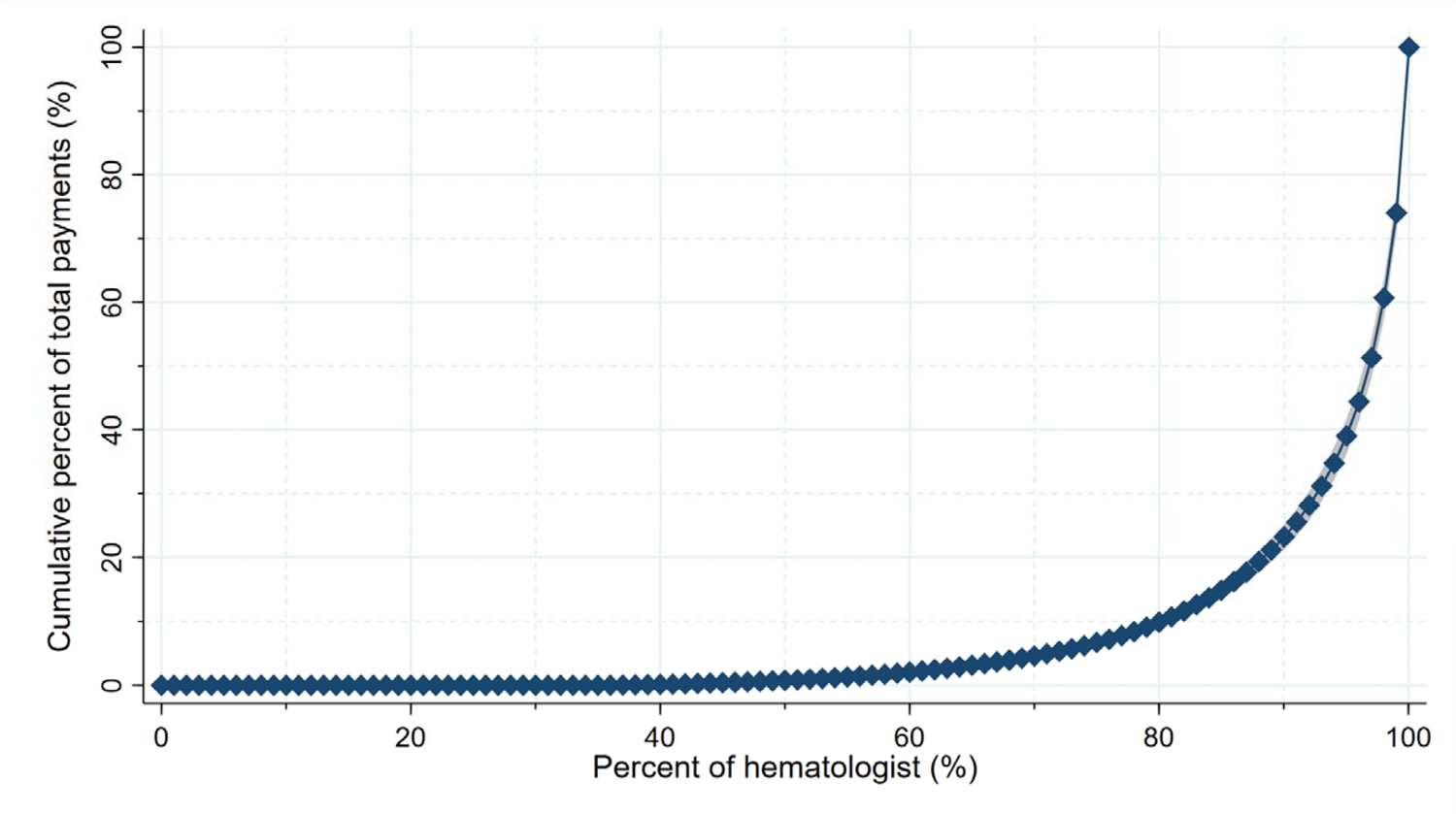
Concentration of payment to individual hematology specialists

### Payment trend between 2016 and 2019

Regarding the payment trend of all 71 companies through the four years, annual total payments increased from $7,700,346 (839,337,686) in 2016 to $10,279,218 (¥1,120,434,752) in 2019. In addition, payments per specialist and the number of specialists with payments increased from $4,259 (SD: $9,291) and 1,808 (43.2%) in 2016 to $5,574 (SD: $11,688) and 1,844 (44.1%) in 2019, respectively. The relative annual increase rates were 1.112 (95%CI: 1.091 1.134; p < 0.001) in payments per specialist and 1.018 (95%CI: 1.006 1.030; p =0.003) for specialists with payments, indicating that the payment values and number of specialists increased by 11.2% (95%CI: 9.1% 13.4%) and 1.8% (95%CI: 0.6% 3.0%) each year (Table 2).

**Table 2.**
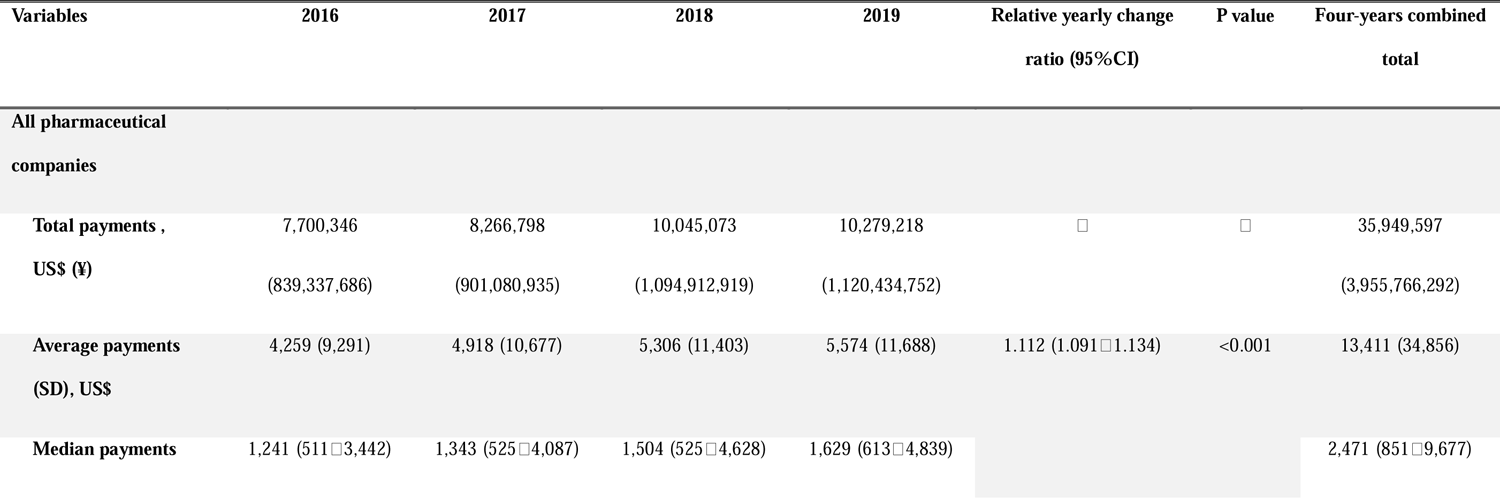
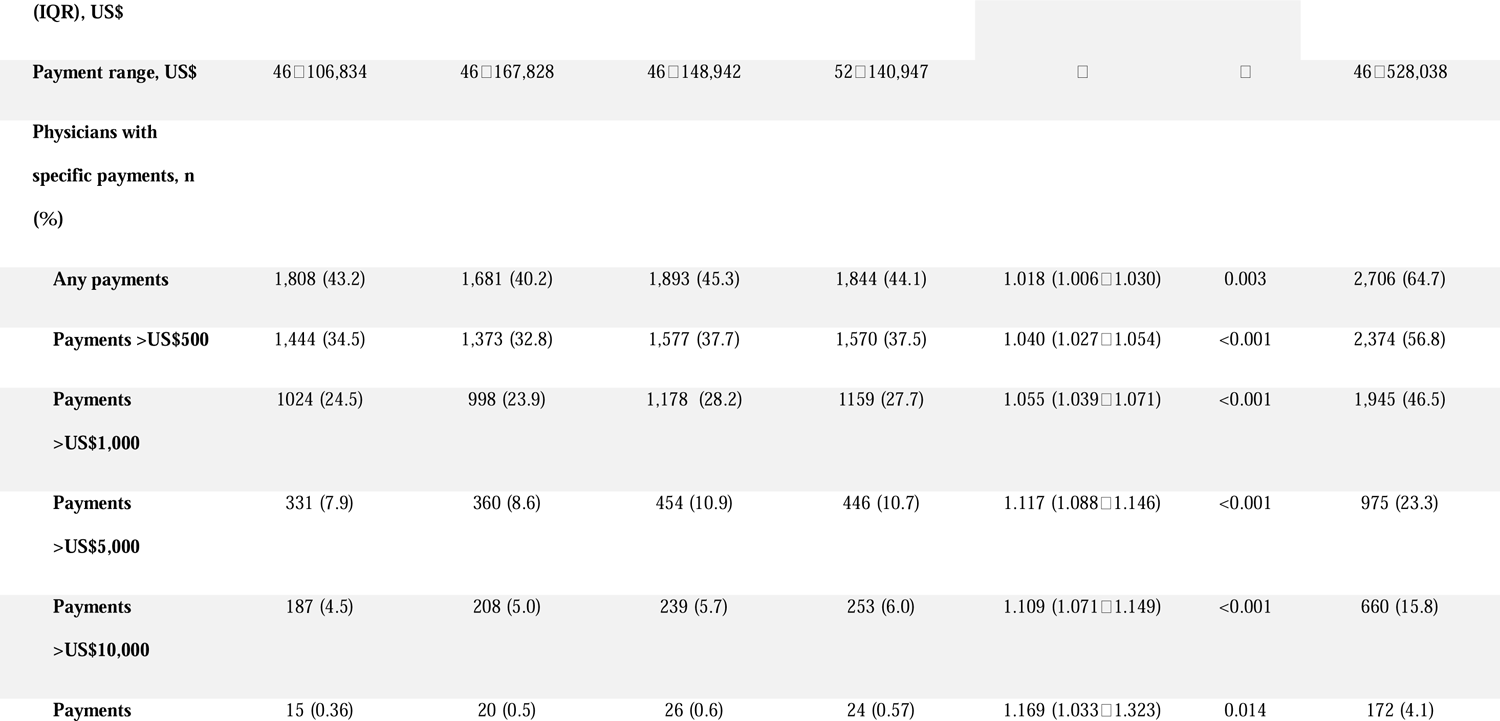
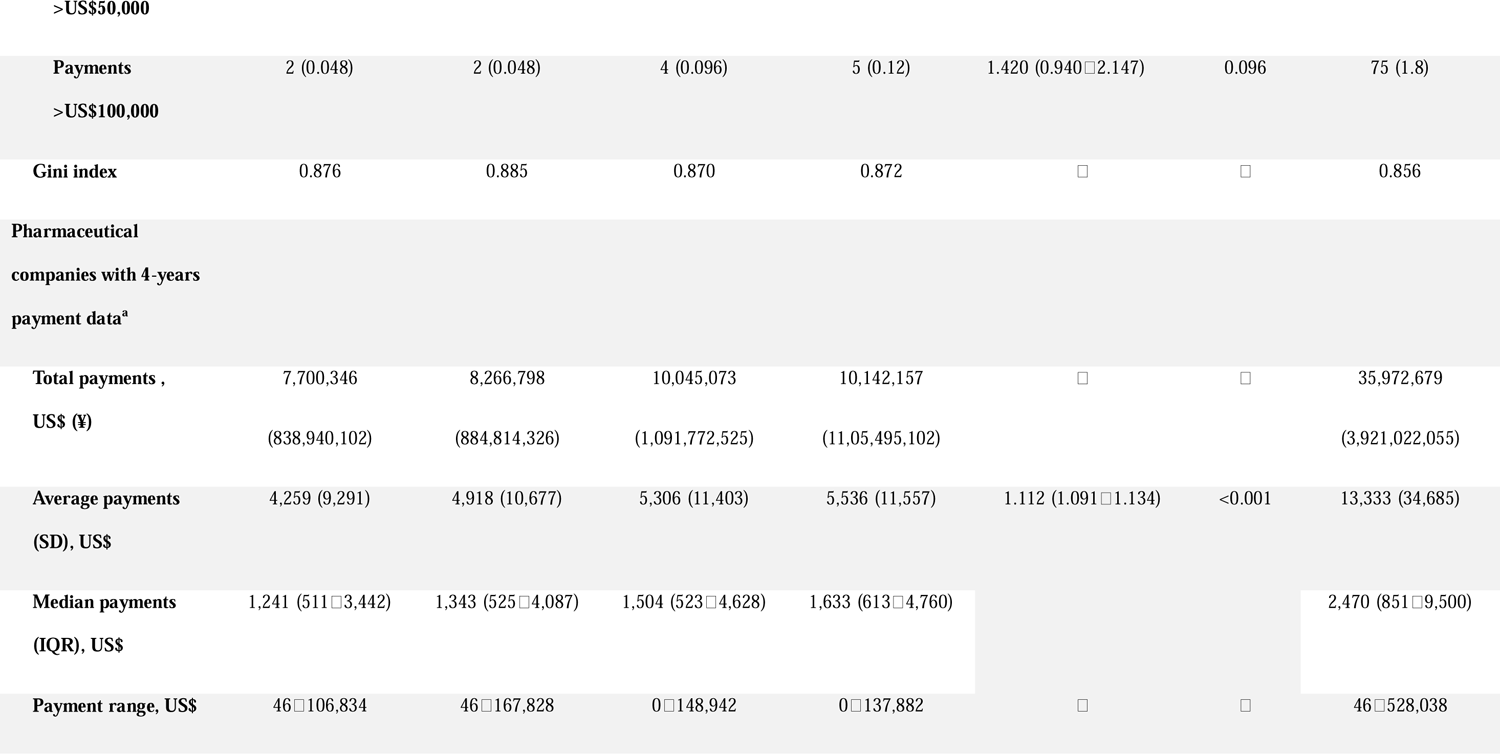
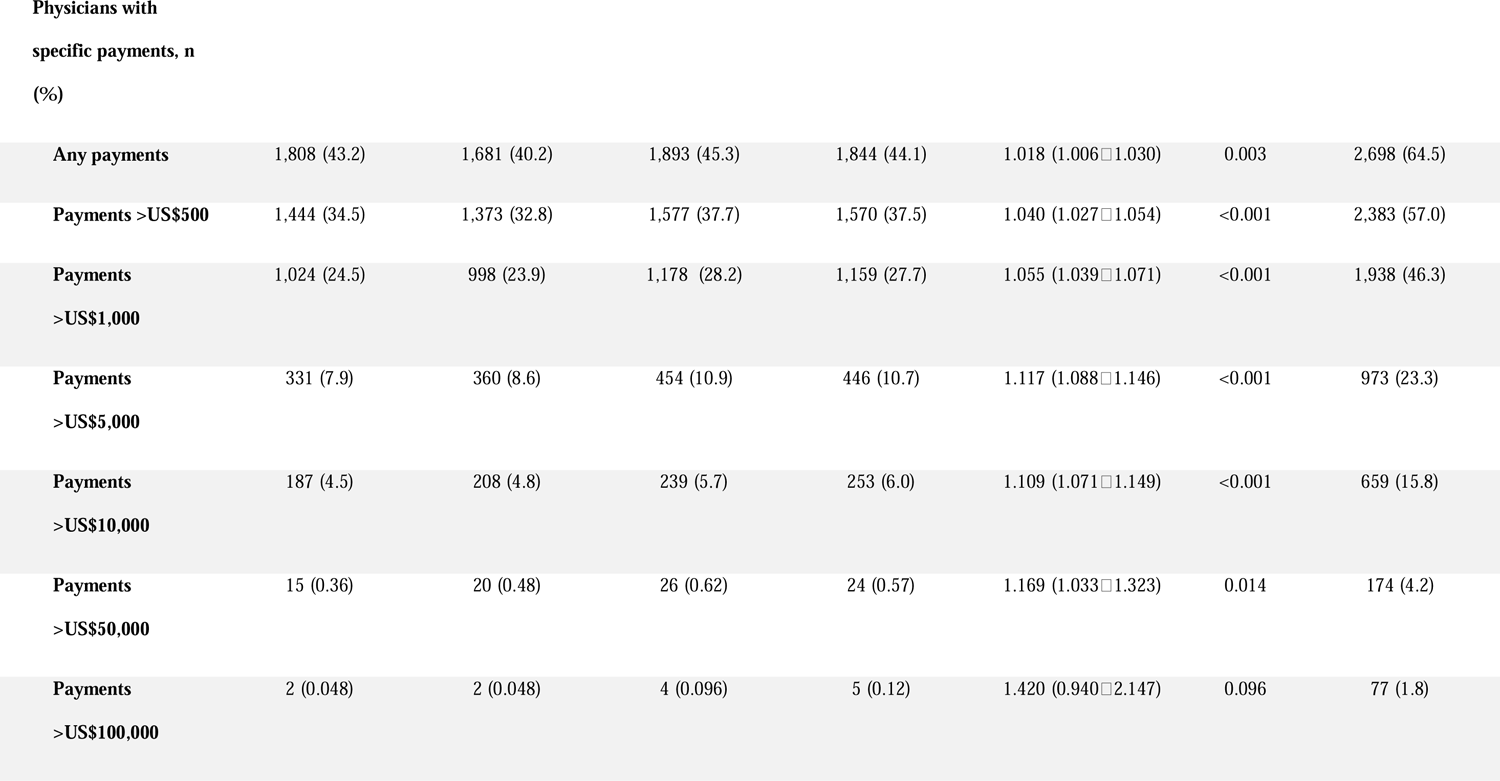
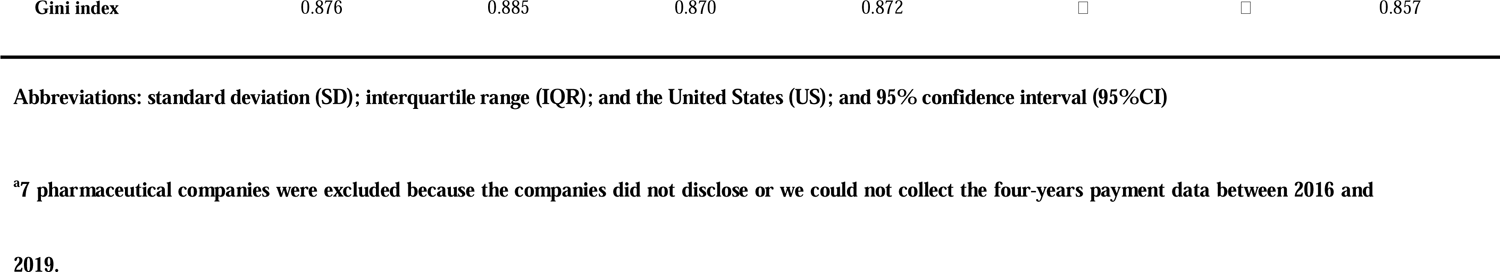
Trend of personal payments from pharmaceutical companies to the hematology specialists board certified by the Japanese Society of Hematology between 2016 and 2019

Of 74 companies collecting payments data over four years, there were 64 companies with the four-years payment data. Limiting to the payments from the 64 companies, both the average and median payment values also constantly increased from $4,259 (SD: $9,291) and $1,241 (IQR: $511 $3,442) to $5,536 (SD: $11,557) and $1,633 (IQR: $613 $4,760) between 2016 and 2019, respectively. The relative annual change rate for payments per specialist and number of specialists with payments also significantly increased by 11.2% (95%CI: 9.1% 13.4%) and 1.8% (95%CI: 0.6% 3.0%), respectively, each year.

Furthermore, the annual change rate was higher among those with greater four-year cumulative payments, such as those receiving $1,000‒$10,000 at 12.8% (95%CI: 9.5% 16.1%; p < 0.001), $10,000‒$50,000 at 13.7% (95%CI: 10.4% 17.1%; p < 0.001), and $50,000‒ $100,000 at 13.8% (95%CI: 8.1% 19.8%; p < 0.001), while those with payments of more than $100,000 had a lower annual increase rate of 8.1% (95%CI: 4.2% 11.1%; p < 0.001).

### Payment by pharmaceutical companies

Among 71 pharmaceutical companies making payments to specialists, payments from the top 10 companies accounted for 70.8% of the total payments ($ 25,236,750) between 2016 and 2019 (Figure 2). The payment types for each of the top 10 paying companies are shown in Figure 3. Four among the 10 companies, namely, Takeda Pharmaceutical, Chugai Pharmaceutical, Janssen Pharmaceutical, and Novartis Pharma, increased their payments to hematology specialists. A total of 673 drugs were newly approved and introduced between 2015 and 2020 in Japan. Among these indicators, 211 (31.4%) were from the top 10 companies, and of these, 52 drugs (24.6%) were for hematological diseases (Supplemental Material 1).

**Figure 2.**
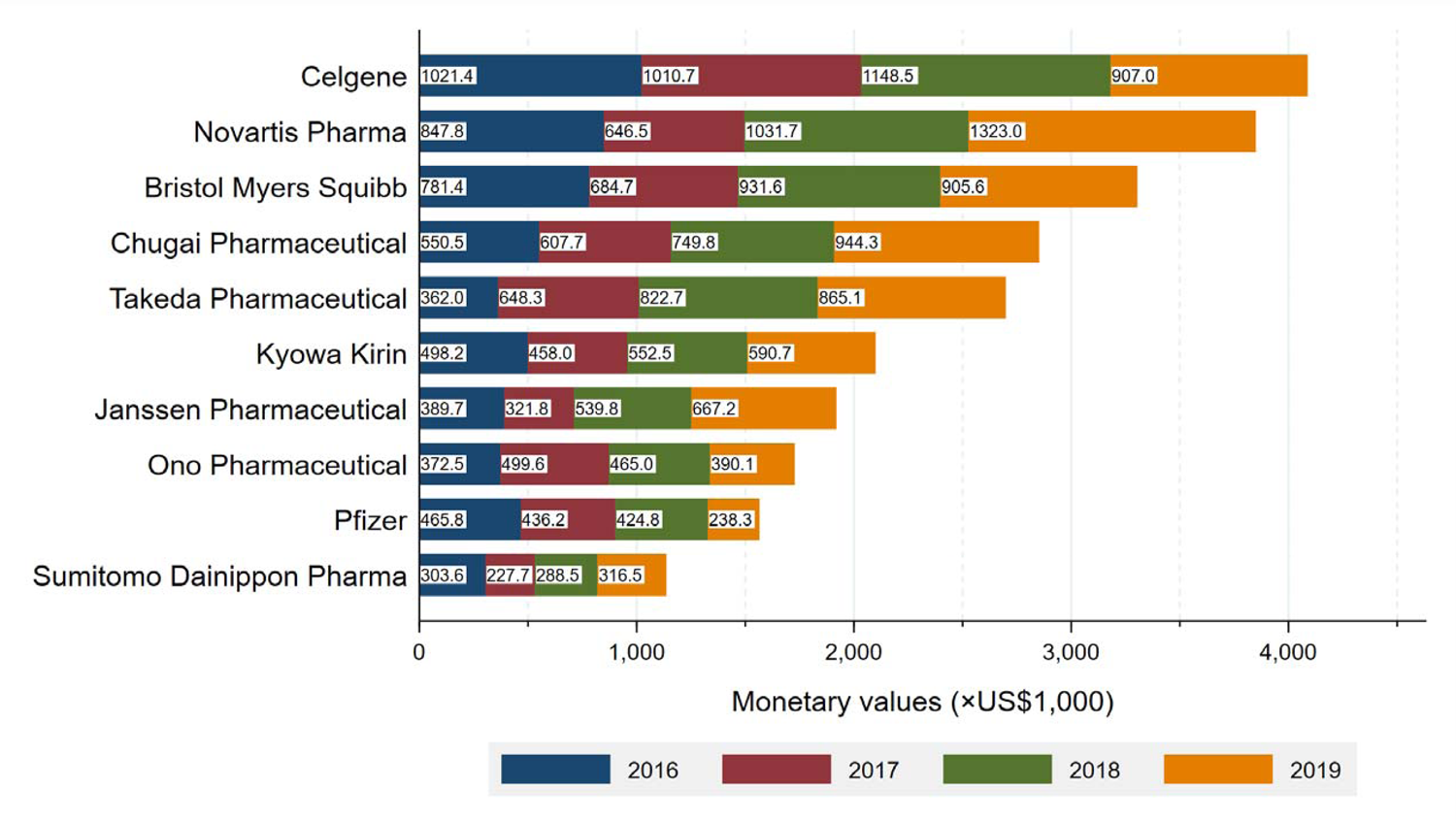
Total payments from top 10 largest paying pharmaceutical companies between 2016 and 2019

**Figure 3.**
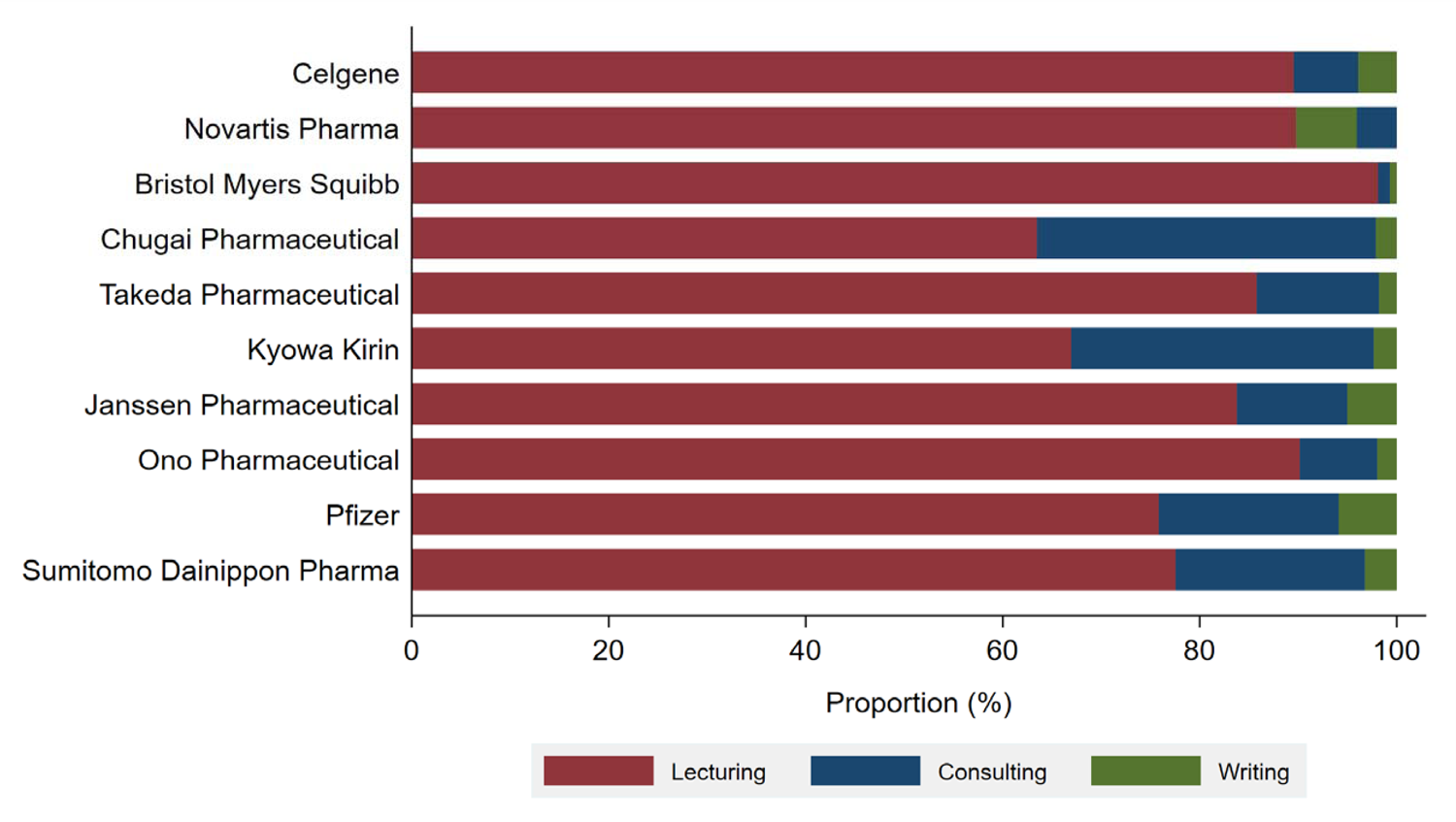
Category and distribution of payments by top 10 largest paying pharmaceutical companies

### Geographical payment distribution

Geographical differences were observed in the distribution of the number of hematology specialists. Based on the prefecture, payments from pharmaceutical companies (Supplemental Material 2A) to hematology specialists per one million of the population ranged from 18.2 in Aomori Prefecture to 64.4 in Kyoto (Supplemental Material 2B). The average payment values per specialist were the highest in Tochigi Prefecture ($15,806) and lowest in Kumamoto Prefecture ($3,178) (Supplemental Material 2D).

## Discussion

This study demonstrated that 3.6% ($ 36,291,434) of total personal payments concerning lecturing, consulting, and writing from all major pharmaceutical companies to healthcare professionals were distributed to board certified hematology specialists, who accounted for 1.3% (4,183 out of 327,210) of total physicians in Japan, according to the latest survey by the Japanese Ministry of Health, Labor and Welfare in 2018(ref.[35]). Among all Japanese board-certified hematology specialists, 64.7% received an average of $13,411 in personal payments between 2016 and 2019, with an annual average increase rate of 11.2 %. To the best of our knowledge, this is the first study to assess the distribution and the trend of financial relationships between board-certified hematologists and pharmaceutical companies. Although our study could have limitations such as underreported payments due to the limited category of personal payments in Japan, there were important similarities and differences between our findings and those of the previous studies.

Previous studies in the United States demonstrated that 80.2% of hematologists and oncologists received general payments averaging $6,166 ($2,055 in one year) between 2015 and 2017. (ref.[31]) Another study by Marshall et al. reported that 63.0% of medical oncologists, including pediatric hematologists/oncologists and hematologist/oncologists, received $7,750 in average general payment in 2014 (ref.[22]), and, in the six years between 2014 and 2019, 84.6% (13,190 out of 15,585) of medical oncologists received $38,601 (6,434 in one year) in average general payment per physician (ref.[20]). Another study by Ozaki et al. evaluated pharmaceutical payments among Japanese board-certified oncologists, and they found that 70.6% of certified oncologists received an average of $4,982 in annual personal payments. (ref.[28]) Our study showed that the average personal payments ranged from $4,259 to $5,574 and the proportion of (mention specialist type) specialists with payments ranging from 40.2% to 45.3% in a single year were fewer than those in the United States and less than the average proportion of Japanese hematologists and oncologists.

Meanwhile, in other specialties, the average total payments per physician, such as general, research, and ownership payments, were $907 among pediatricians in 2014 (ref.[36]); $4,183 ($2,091 per year) among psychiatrists between 2016 and 2017 (ref.[37]); $3,433 ($1,144 in one year) in general payments among obstetricians and gynecologists between 2013 and 2015 (ref.[38]); from $1,795 in 2014 to $2,227 in 2017 among board certificated nephrologists (ref.[39]); $3,260 among orthopedic surgeons in 2013 (ref.[40]); $4,177 among dermatologists in 2014 (ref.[30]); $25,960 ($5,191 per year) among ophthalmologists between 2013 and 2017 (ref.[41]); $16,914 ($5,638 per year) among cardiologists between 2014 and 2016 (ref.[42]); $38,664 ($6,444 per year) among rheumatologists between 2014 and 2019 (ref.[43]); and $7,263 among neurologists in 2015. (ref.[44]) Comparing these findings, the Japanese board certificated hematologists might have similar or greater financial relationships with pharmaceutical companies than other specialists in the United States and Japan. This study showed that only a very small proportion of hematology specialists received the majority of personal payments from pharmaceutical companies. As previous studies have found (ref.[25, 30, 41–43]), pharmaceutical companies have concentrated their payments on a small number of physicians with extensive clinical and research experience. These specialists, or “key opinion leaders,” (ref.[45]) who had substantial financial relationships with pharmaceutical companies, played a significant role in delivering information about newer drugs, novel findings from clinical trials, and drug related risks, to healthcare professionals at conferences for continuing medical education sponsored by pharmaceutical companies (ref.[46]). We should note that while the financial relationships of specialists with pharmaceutical companies might help to rapidly unroll novel drugs with greater benefits (ref.[26, 46]), it could also increase healthcare costs when the specialists prescribe brand name drugs (ref.[4, 47]) or drugs whose benefits are unproven (ref.[48, 49]).

Further, regarding the trend of payments, we found that total and average payments increased significantly every year, with an 11.2% yearly increase in average payments. This trend of increasing payments to physicians was also observed among oncologists in the United States. Marshall et al. found that since the launch of the US Open Payment Database, the total and annual average payment value per physician declined by −1.7% and −0.6%, respectively (ref.[34]). However, pharmaceutical companies increasingly prioritized the payments to hematologists and oncologists, with a 4.9% and 1.7% annual increase in total value and average payments (ref.[20]). Furthermore, although the disclosure of personal payments from pharmaceutical companies to healthcare professionals and healthcare organizations was intended to improve transparency rather than curb the financial relationships, our findings indicate that disclosure itself did not sufficiently decrease the financial relationships between pharmaceutical companies and hematology specialists in Japan, as corroborated by previous studies in the United States.

Companies’ payment trends would provide further understanding of the increase in the average payments and proportion of specialists with payments among Japanese hematology specialists. The top 10 companies expanded their indications in the field of hematology, ranging from one to 11 new indications per company. While the payment from Celgene, the largest paying company, remained stable, four companies, namely, Takeda Pharmaceutical, Chugai Pharmaceutical, Janssen Pharmaceutical, and Novartis Pharma, remarkably increased their payments to the hematology specialists between 2016 and 2019. As a drug target, multiple myeloma accounted for the largest proportion (17 out of 52 indications), which may explain the recent trend of increasing payments. In fact, all the companies mentioned above, except Chugai Pharmaceutical, have developed newly approved drugs for multiple myeloma. In addition to multiple myeloma, Novartis made the largest total payments of $1,323,027 in 2019, when the chimeric antigen receptor T cells, tisagenlecleucel (KYMRIAH), was granted approval the treatment of B-cell precursor acute lymphoblastic leukemia and large B-cell lymphoma and was priced at about 32 million JPY (equal to about 300,000 USD), covered by the Japanese public health insurance. Regarding Chugai Pharmaceutical, emicizumab (HEMLIBRA, approved in 2018 for congenital blood coagulation factor 8 deficiency) may account for the increased payment.

This study has several limitations. First, our payment database was constructed by manually collecting payment data from 92 pharmaceutical companies and repeatedly crosschecking it for any errors by two or more persons. Despite careful and repeated checks, the inclusion of errors by our study team and pharmaceutical companies reporting data could not be ruled out. However, the publicly disclosed payment database (https://db.tansajp.org/en) welcomed inquiries from the recipients of payments and pharmaceutical companies reporting any errors in the data, and the payment database was checked and revised for each inquiry. These procedures minimized the inclusion of errors in this study. In addition, payment data concerning meals, education, travel, and accommodations were not disclosed with the individual names of recipients by pharmaceutical companies in Japan (ref.[16]). Thus, these payment data are not analyzable. Considering that such payment categories like meals were the most prevalent among physicians (ref.[30, 42, 48]), this study underreports the prevalence and magnitude of financial relationships between hematology specialists and pharmaceutical companies in Japan. However, this study highlights the magnitude and prevalence of financial relationships between pharmaceutical companies and Japanese hematologists.

In conclusion, this study found that between 2016 and 2019, 64.7% of Japanese board-certified hematology specialists received an average of $13,411 in personal payments as the reimbursement for lecturing, consulting, and writing. Only a small proportion of hematology specialists received the vast majority of payments from pharmaceutical companies. Furthermore, these personal payments from pharmaceutical companies were increasingly more prevalent and greater among Japanese hematology specialists.

## Data Availability

All data produced in the present study are available upon reasonable request to the authors.

## Acknowledgement

The authors thank the Tansa (formerly known as Waseda Chronicle) for providing payment data and Editage (www.editage.com) for English language editing. Also, we appreciate Mr. Souto Nagano, an undergraduate student at the University of Tokyo Faculty of Letters; Mr. Kohki Yamada, a medical student at the Osaka University School of Medicine; and Ms. Megumi Aizawa, a graduate student at the Department of Industrial Engineering and Economics, School of Engineering, Tokyo Institute of Technology, for their dedicated contribution on collecting and cross-checking the payment data.

This study was funded in part by the Medical Governance Research Institute. This non-profit enterprise receives donations from pharmaceutical companies, including Ain Pharmacies, Inc., other organizations, and private individuals. This study also received support from the Tansa (formerly known as the Waseda Chronicle), an independent non-profit news organization dedicated to investigative journalism. None of the entities providing financial support for this study contributed to the design, execution, data analyses, or interpretation of study findings and the drafting of this manuscript.

For the financial conflicts of interest, Dr. Kusumi received personal fees from Otsuka Pharmaceutical Co. Ltd outside the scope of the submitted work. Dr. Saito received personal fees from TAIHO Pharmaceutical Co. Ltd outside the scope of the submitted work. Drs. Ozaki and Tanimoto received personal fees from Medical Network Systems outside the scope of the submitted work. Dr. Tanimoto also received personal fees from Bionics Co. Ltd, outside the scope of the submitted work. Regarding non-financial conflicts of interest among the study authors, all are engaged in ongoing research examining financial and non-financial conflicts of interest among healthcare professionals and pharmaceutical companies in Japan. Individually, Anju Murayama, Hiroaki Saito, Toyoaki Sawano, Tetsuya Tanimoto, and Akihiko Ozaki have contributed to several published studies assessing conflicts of interest and quality of evidence among clinical practice guideline authors in Japan and the United States. Among their previous articles, the authors have self-cited several articles in this study to gain deeper insights and explain the context of financial conflicts of interest among healthcare professionals in Japan. Dr. Kusumi was a hematology specialist board certificated by the Japanese Society of Hematology. The other authors have no example conflicts of interest to disclose.

## Author Contributions

EK was responsible for study concept, data collection, statistical analysis, and drafting and reviewing of the manuscript. AM was responsible for study concept and design, data collection, statistical analysis, and drafting and reviewing of the manuscript. SK was responsible for study concept and design, data collection, statistical analysis, and drafting of the manuscript. MK contributed to study concept, data collection, and drafting of the manuscript. MY contributed to study concept, data collection, and drafting of the manuscript. HS contributed to study concept and design, drafting of the manuscript, and critically reviewing of the manuscript. TS contributed to study concept and design, drafting of the manuscript, and critically reviewing of the manuscript. EY contributed to study concept and design, and critically reviewing of the manuscript. TT conducted to study concept and design, drafting of the manuscript, and study supervision. AO conducted study concept and design, statistical analysis, drafting of the manuscript, and study supervision. All authors had full access to all the data in the study and take responsibility for the integrity of the data and the accuracy of the data analysis.

## References

1. Wouters OJ, McKee M, Luyten J. Estimated Research and Development Investment Needed to Bring a New Medicine to Market, 2009-2018. JAMA 2020;323(9):844-853.

2. Rubin EH, Gilliland DG. Drug development and clinical trials—the path to an approved cancer drug. Nature Reviews Clinical Oncology 2012;9(4):215–222.

3. Medicine AFABoI, Medicine A-AFACoP-ASoI, European Federation of Internal M. Medical professionalism in the new millennium: a physician charter. Ann Intern Med 2002;136(3):243–6.

4. DeJong C, Aguilar T, Tseng C-W, et al. Pharmaceutical Industry–Sponsored Meals and Physician Prescribing Patterns for Medicare Beneficiaries. JAMA Internal Medicine 2016;176(8):1114–1122.

5. Mitchell AP, Trivedi NU, Gennarelli RL, et al. Are Financial Payments From the Pharmaceutical Industry Associated With Physician Prescribing?: A Systematic Review. Ann Intern Med 2020; 10.7326/m20-5665.

6. Hartung DM, Johnston K, Cohen DM, et al. Industry Payments to Physician Specialists Who Prescribe Repository Corticotropin. JAMA Network Open 2018;1(2):e180482–e180482.

7. Nejstgaard CH, Bero L, Hrobjartsson A, et al. Association between conflicts of interest and favourable recommendations in clinical guidelines, advisory committee reports, opinion pieces, and narrative reviews: systematic review. BMJ 2020;371:m4234.

8. Murayama A, Ozaki A, Saito H, et al. Are Clinical Practice Guideline for Hepatitis C by the American Association for the Study of Liver Disease and Infectious Diseases Society of America Evidence-based? Financial Conflicts of Interest and Assessment of Quality of Evidence and Strength of Recommendations. Hepatology 2021;n/a(n/a).

9. Johnson L, Stricker RB. Attorney General forces Infectious Diseases Society of America to redo Lyme guidelines due to flawed development process. J Med Ethics 2009;35(5):283–8.

10. Coyne DW. Influence of Industry on Renal Guideline Development. Clinical Journal of the American Society of Nephrology 2007;2(1):3–7.

11. Hashimoto T, Murayama A, Mamada H, et al. Evaluation of Financial Conflicts of Interest and Drug Statements in the Coronavirus Disease 2019 Clinical Practice Guideline in Japan. Clin Microbiol Infect 2021; 10.1016/j.cmi.2021.11.019.

12. Lundh A, Lexchin J, Mintzes B, et al. Industry sponsorship and research outcome. Cochrane Database Syst Rev 2017;2(2):MR000033.

13. Wilson JM. Lessons learned from the gene therapy trial for ornithine transcarbamylase deficiency. Mol Genet Metab 2009;96(4):151–7.

14. Pham-Kanter G. Act II of the Sunshine Act. PLoS Med 2014;11(11):e1001754.

15. Silverman E. Everything you need to know about the Sunshine Act. BMJ: British Medical Journal 2013;347:f4704.

16. Ozaki A, Saito H, Senoo Y, et al. Overview and transparency of non-research payments to healthcare organizations and healthcare professionals from pharmaceutical companies in Japan: Analysis of payment data in 2016. Health Policy 2020;124(7):727–735.

17. Zhou J, Vallejo J, Kluetz P, et al. Overview of Oncology and Hematology Drug Approvals at US Food and Drug Administration Between 2008 and 2016. JNCI: Journal of the National Cancer Institute 2019;111(5):449–458.

18. Mato AR, Shah NN, Jurczak W, et al. Pirtobrutinib in relapsed or refractory B-cell malignancies (BRUIN): a phase 1/2 study. Lancet 2021;397(10277):892–901.

19. Awasthi R, Pacaud L, Waldron E, et al. Tisagenlecleucel cellular kinetics, dose, and immunogenicity in relation to clinical factors in relapsed/refractory DLBCL. Blood Advances 2020;4(3):560–572.

20. Tarras ES, Marshall DC, Rosenzweig K, et al. Trends in Industry Payments to Medical Oncologists in the United States Since the Inception of the Open Payments Program, 2014 to 2019. JAMA Oncology 2021;7(3):440-444.

21. Pokorny AMJ, Bero LA, Moynihan R, et al. Industry payments to Australian medical oncologists and clinical haematologists: a cross-sectional analysis of publicly-available disclosures. Intern Med J 2020;n/a(n/a).

22. Marshall DC, Moy B, Jackson ME, et al. Distribution and Patterns of Industry-Related Payments to Oncologists in 2014. JNCI: Journal of the National Cancer Institute 2016;108(12).

23. Harada K, Ozaki A, Saito H, et al. Financial payments made by pharmaceutical companies to the authors of Japanese hematology clinical practice guidelines between 2016 and 2017. Health Policy 2021;125(3):320–326.

24. Murayama A, Ozaki A, Saito H, et al. Pharmaceutical company payments to dermatology Clinical Practice Guideline authors in Japan. PLoS One 2020;15(10):e0239610.

25. Yamamoto K, Murayama A, Ozaki A, et al. Financial conflicts of interest between pharmaceutical companies and the authors of urology clinical practice guidelines in Japan. Int Urogynecol J 2021;32(2):443–451.

26. Mitchell AP, Winn AN, Dusetzina SB. Pharmaceutical Industry Payments and Oncologists’ Selection of Targeted Cancer Therapies in Medicare Beneficiaries. JAMA Internal Medicine 2018;178(6):854–856.

27. Mitchell AP, Winn AN, Lund JL, et al. Evaluating the Strength of the Association Between Industry Payments and Prescribing Practices in Oncology. The Oncologist 2019;24(5):632–639.

28. Ozaki A, Saito H, Onoue Y, et al. Pharmaceutical payments to certified oncology specialists in Japan in 2016: a retrospective observational cross-sectional analysis. BMJ Open 2019;9(9):e028805.

29. Tringale KR, Marshall D, Mackey TK, et al. Types and Distribution of Payments From Industry to Physicians in 2015. JAMA 2017;317(17):1774–1784.

30. Feng H, Wu P, Leger M. Exploring the Industry-Dermatologist Financial Relationship: Insight From the Open Payment Data. JAMA Dermatol 2016;152(12):1307–1313.

31. Inoue K, Blumenthal DM, Elashoff D, et al. Association between physician characteristics and payments from industry in 2015-2017: observational study. BMJ Open 2019;9(9):e031010.

32. Tringale KR, Hattangadi-Gluth JA. Types and Distributions of Biomedical Industry Payments to Men and Women Physicians by Specialty, 2015. JAMA Intern Med 2018;178(3):421-423.

33. Marshall DC, Tarras ES, Rosenzweig K, et al. Trends in Financial Relationships Between Industry and Radiation Oncologists Versus Other Physicians in the United States from 2014 to 2018. International Journal of Radiation Oncology*Biology*Physics 2021;109(1):15–25.

34. Marshall DC, Tarras ES, Rosenzweig K, et al. Trends in Industry Payments to Physicians in the United States From 2014 to 2018. JAMA 2020;324(17):1785–1788.

35. Ministry of Health LaW. Survey of Physicians, Dentists and Pharmacists 2018. In: Ministry of Health, Labour and Welfare; 2018.

36. Parikh K, Fleischman W, Agrawal S. Industry Relationships With Pediatricians: Findings From the Open Payments Sunshine Act. Pediatrics 2016;137(6):e20154440.

37. Rhee TG, Wilkinson ST. Exploring the Psychiatrist-Industry Financial Relationship: Insight from the Open Payment Data of Centers for Medicare and Medicaid Services. Adm Policy Ment Health 2020;47(4):526–530.

38. Muffly TM, Giamberardino WL, Guido J, et al. Industry Payments to Obstetricians and Gynecologists Under the Sunshine Act. Obstetrics & Gynecology 2018;132(1).

39. Pakanati AR, Kovvuru K, Thombre V, et al. Industry Payments to Nephrologists in the United States. Cureus 2021;13(8):e17057.

40. Cvetanovich GL, Chalmers PN, Bach BR, Jr. Industry Financial Relationships in Orthopaedic Surgery: Analysis of the Sunshine Act Open Payments Database and Comparison with Other Surgical Subspecialties. JBJS 2015;97(15).

41. Slentz DH, Nelson CC, Lichter PR. Characteristics of Industry Payments to Ophthalmologists in the Open Payments Database. JAMA Ophthalmology 2019;137(9):1038–1044.

42. Annapureddy A, Murugiah K, Minges KE, et al. Industry Payments to Cardiologists. Circulation: Cardiovascular Quality and Outcomes 2018;11(12):e005016.

43. Putman MS, Goldsher JE, Crowson CS, et al. Industry Payments to Practicing US Rheumatologists, 2014–2019. Arthritis & Rheumatology 2021;73(11):2138-2144.

44. Ahlawat A, Narayanaswami P. Financial relationships between neurologists and industry. Neurology 2019;92(21):1006.

45. Moynihan R. Key opinion leaders: independent experts or drug representatives in disguise? BMJ 2008;336(7658):1402-3.

46. Inoue K, Figueroa JF, DeJong C, et al. Association Between Industry Marketing Payments and Prescriptions for PCSK9 (Proprotein Convertase Subtilisin/Kexin Type 9) Inhibitors in the United States. Circulation: Cardiovascular Quality and Outcomes 2021;14(5):e007521.

47. Yeh JS, Franklin JM, Avorn J, et al. Association of Industry Payments to Physicians With the Prescribing of Brand-name Statins in Massachusetts. JAMA Internal Medicine 2016;176(6):763–768.

48. Inoue K, Tsugawa Y, Mangione CM, et al. Association between industry payments and prescriptions of long-acting insulin: An observational study with propensity score matching. PLOS Medicine 2021;18(6):e1003645.

49. Goupil B, Balusson F, Naudet F, et al. Association between gifts from pharmaceutical companies to French general practitioners and their drug prescribing patterns in 2016: retrospective study using the French Transparency in Healthcare and National Health Data System databases. BMJ 2019;367:l6015.

