## Supplemental Materials 1&2 for "Pharmaceutical Payments to Japanese Certificated Hematologists: A Retrospective Analysis of Personal Payments from Pharmaceutical Companies between 2016 and 2019"

**Supplemental Material 1. Newly approved drugs for hematological disease between 2015 and 2020 in Japan**

| Company | General name | Brand name | Approval date | Category of approval | Type of disease |
| --- | --- | --- | --- | --- | --- |
| Celgene | Lenalidomide hydrate | Revlimid | February 21, 2020 | Added indication | Follicular lymphoma  Marginal zone lymphoma |
|  | Pomalidomide | Pomalyst | May 22, 2019 | Added indication | Multiple myeloma |
|  | Romidepsin | Istodax | July 3, 2017 | New approval | Peripheral T-cell lymphoma |
|  | Lenalidomide hydrate | Revlimid | March 2, 2017 | Added indication | Adult T-cell leukemia |
|  | Lenalidomide hydrate | Revlimid | December 21, 2015 | Added indication | Multiple myeloma |
|  | Pomalidomide | Pomalyst | March 26, 2015 | New approval | Multiple myeloma |
| Novartis Pharma | Tisagenlecleucel | Kymriah | March 26, 2019 | New approval | CD19-positive B-cell acute lymphoblastic leukemia  CD19-positive diffuse large B-cell lymphoma |
|  | Nilotinib Hydrochloride Hydrate | Tasigna | December 25, 2017 | Added indication | Chronic myelogenous leukemia |
|  | Eltrombopag olamine | Revolade | August 25, 2017 | Added indication | Aplastic anemia |
|  | Cyclosporine | Neoral | August 25, 2017 | Added indication | Aplastic anemia |
|  | Deferasirox | Jadenu | July 3, 2017 | New approval | Chronic iron overload |
|  | Ruxolitinib Phosphate | Jakavi | September 24, 2015 | Added indication | Polycythemia vera |
|  | Panobinostat lactate | Farydak | July 3, 2015 | New approval | Multiple myeloma |
| Takeda Pharmaceutical | Vonicog alfa^a^ | Vonvendi | March 25, 2020 | New approval | Von willebrand disease |
|  | Ixazomibe citrate ester | Ninlaro | March 25, 2020 | Added indication | Multiple myeloma |
|  | Brentuximab vedotin | Adcetris | September 21, 2018 | Added indication | CD30-positive hodgkin lymphoma |
|  | Rurioctocog alfa pegol^b^ | Adynovate | November 30, 2017 | Added indication | Congenital blood coagulation factor 8 deficiency |
|  | Ixazomibe citrate ester | Ninlaro | March 30, 2017 | New approval | Multiple myeloma |
|  | Prednisolone | Prednisolone | June 26, 2015 | Added indication | Multiple myeloma |
| Bristol Myers Squibb | Elotuzumab | Empliciti | November 22, 2019 | Added indication | Multiple myeloma |
|  | Elotuzumab | Empliciti | September 28, 2016 | New approval | Multiple myeloma |
| Chugai Pharmaceutical | Alectinib hydrochloride | Alecensa | February 21, 2020 | Added indication | Anaplastic large cell lymphoma |
|  | Emicizumab | Hemlibra | December 21, 2018 | Added indication | Congenital blood coagulation factor 8 deficiency with inhibitor against blood coagulation factor 8 |
|  | Emicizumab | Hemlibra | December 21, 2018 | Added indication | Congenital blood coagulation factor 8 deficiency without inhibitor against blood coagulation factor 8 |
|  | Obinutuzumab | Gazyva | July 2, 2018 | New approval | CD20-positive follicular lymphoma |
|  | Emicizumab | Hemlibra | March 23, 2018 | New approval | Congenital blood coagulation factor 8 deficiency |
| Kyowa Kirin | Romiplostim | Romiplate | June 18, 2019 | Added indication | Aplastic anemia |
|  | Mogamulizumab | Poteligeo | August 21, 2018 | Added indication | CCR4-positive adult T-cell leukemia-lymphoma  CCR4-positive peripheral t-cell lymphoma |
|  | Antithrombin gamma | Acoalan | July 3, 2015 | New approval | Congenital antithrombin deficiency |
| Janssen Pharmaceutical | Daratumumab | Darzalex | November 27, 2020 | Added indication | Multiple myeloma |
|  | Daratumumab | Darzalex | December 20, 2019 | Added indication | Multiple myeloma |
|  | Daratumumab | Darzalex | August 22, 2019 | Added indication | Multiple myeloma |
|  | Bortezomib | Velcade | August 22, 2019 | Added indication | Multiple myeloma |
|  | Ibrutinib | Imbruvica | July 2, 2018 | Added indication | Chronic lymphocytic leukemia |
|  | Bortezomib | Velcade | March23, 2018 | Added indication | Primary macroglobulinemia  Lymphatic plasma cell lymphoma |
|  | Daratumumab | Darzalex | September 27, 2017 | New approval | Multiple myeloma |
|  | Ibrutinib | Imbruvica | December 2, 2016 | Added indication | Mantle cell lymphoma |
|  | Ibrutinib | Imbruvica | March 28, 2016 | New approval | Chronic lymphocytic leukemia |
|  | Bortezomib | Velcade | June 26, 2015 | Added indication | Mantle cell lymphoma |
| Ono Pharmaceutical | Nivolumab | Opdivo | September 25, 2020 | Added indication | Classic Hodgkin lymphoma |
|  | Tirabrutinib hydrochloride | Velexbru | August 21, 2020 | Added indication | Primary macroglobulinemia  Lymphatic plasma cell lymphoma |
|  | Carfilzomib | Kyprolis | November 22, 2019 | Added indication | Multiple myeloma |
|  | Nivolumab | Opdivo | September 21, 2018 | New approval | Classic Hodgkin lymphoma |
|  | Nivolumab | Opdivo | August 21, 2018 | Added indication | Classic Hodgkin lymphoma |
|  | Carfilzomib | Kyprolis | May 18, 2017 | Added indication | Multiple myeloma |
|  | Nivolumab | Opdivo | December 2, 2016 | Added indication | Classic Hodgkin lymphoma |
|  | Carfilzomib | Kyprolis | July 4, 2016 | New approval | Multiple myeloma |
| Pfizer | Bosutinib hydrate | Bosulif | June 29, 2020 | Added indication | Chronic myelogenous leukemia |
|  | Rituximab | Rituximab biosimilar | September 20, 2019 | New approval | CD20-positive B-cell non-Hodgkin lymphoma  CD20-positive B-cell lymphoproliferative disorder |
|  | Inotuzumab ozogamicin | Besponsa | January 19, 2018 | New approval | CD22-positive acute lymphocytic leukemia |
|  | Voriconazole | Vfend | August 24, 2015 | Added indication | Prevention of cryptosporidiosis for hematopoietic stem cell |
| Sumitomo Dainippon Pharma | Thiotepa | Rethio | March 25, 2020 | Added indication | Malignant lymphoma |

^a^Vonicog alfa (Vonvendi) was developed by Shire Japan, but due to the merger between Takeda Pharmaceutical and Shire Japan in 2019, we included vonicog alfaa as a product from Takeda Pharmaceutical. ^b^Rurioctocog alfa pegol was developed by Baxalta, and due to the merger between Baxalta and Shire Japan in 2016 and between Takeda Pharmaceutical and Shire Japan in 2019, we included vonicog alfaa as a product from Takeda Pharmaceutical.

**Supplemental Material 2. Geographical characteristics of specialist and payment distribution**
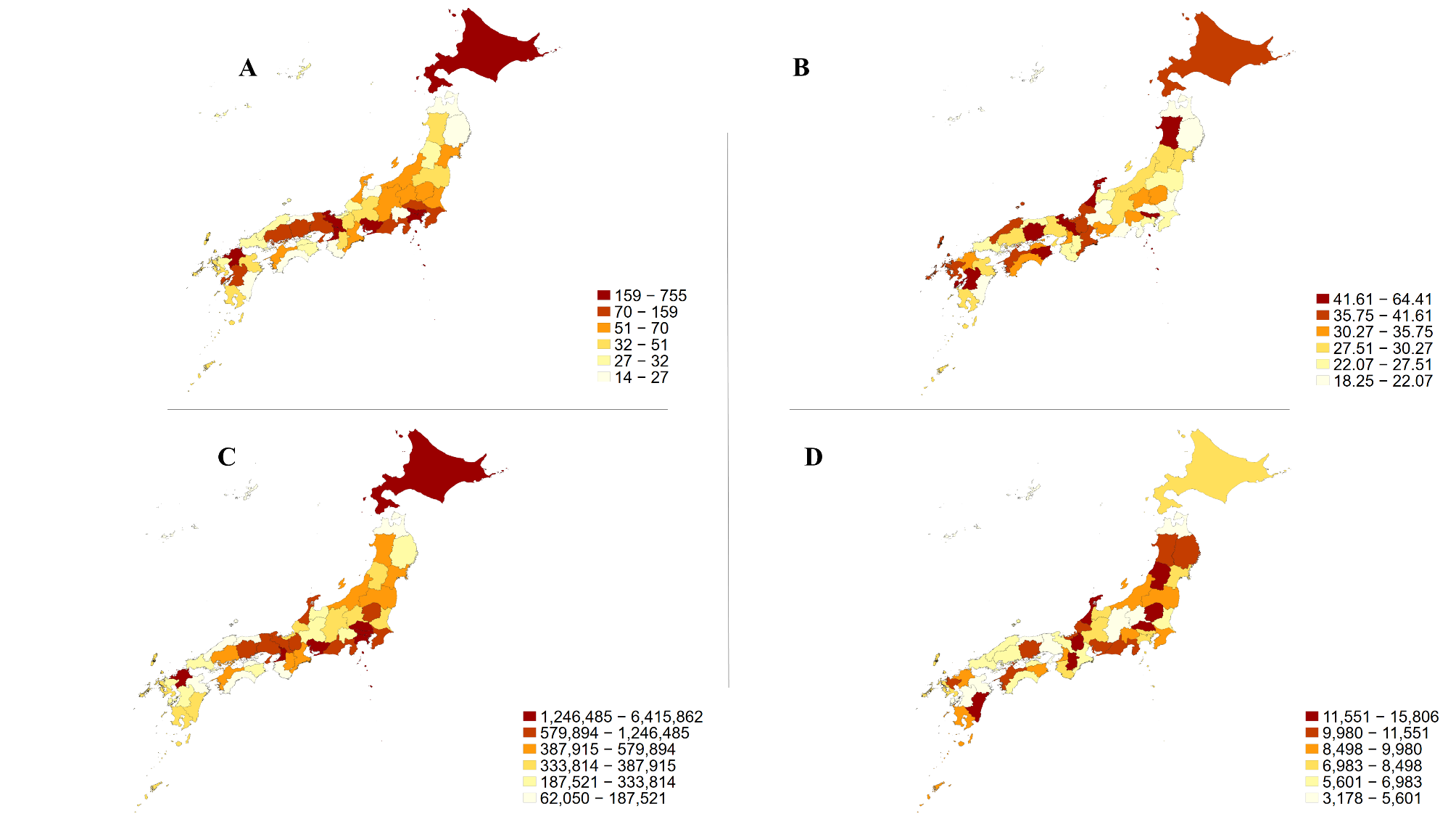

Number of hematology specialists in 2021(A); number of hematology specialists per one million population in 2021(B); total personal payments from pharmaceutical companies to the Japanese hematology specialists over the four-year period 2016 to 2019(C); and total personal payments from pharmaceutical companies to the Japanese hematology specialists per one million population over the four-year period 2016 to 2019(D)
